# Variability in peroneus brevis tendon position on magnetic resonance at the lateral malleolus: an observational cohort study

**DOI:** 10.1101/2025.05.18.25327851

**Authors:** Rafał Zych, Dawid Dziedzic, Katarzyna Bokwa-Dąbrowska, Dan Mocanu, Pawel Szaro

**Affiliations:** Department of Clinical and Descriptive Anatomy, Medical University of Warsaw, Poland; Department of Radiology, Institute of Clinical Sciences, Sahlgrenska Academy, University of Gothenburg, Gothenburg, Sweden; Department of Musculoskeletal Radiology, Sahlgrenska University Hospital, Gothenburg, Sweden

## Abstract

**Purpose:** This study aimed to classify anatomical variants in peroneus brevis position and assess associations with tendon shape, size, and the presence of the peroneus quartus muscle and low-lying peroneus brevis muscle.

**Methods:** This observational cohort study included 230 ankle magnetic resonance examinations (3T) with normal peroneal tendons. Peroneus brevis position relative to peroneus longus was categorized into four types: medial, overlapping with medial protrusion, overlapping with lateral protrusion, and overlapping with both. Tendon shape was classified as general flat, flattened convex medially, flattened convex laterally, or oval. Associations between position and shape were tested using chi-square. Differences in cross-sectional area (mm²) and width (mm) across groups were assessed with analysis of variance and Tukey’s post hoc test. A regression model identified predictors of tendon overlap.

**Results:** The most common position was overlap with medial protrusion (72.0%), followed by medial, lateral, and combined protrusions. Position was significantly associated with shape (p < 0.001); oval tendons were typically medial, while flattened tendons overlapped. Width and cross-sectional area differed significantly across positions (p = 0.0088), with the largest area in tendons protruding medially and laterally (16.9 mm²). Width correlated strongly with overlap (r = 0.79, p <0.001) and was the strongest predictor in regression (β = 0.51, p <0.001). Peroneus quartus was independently associated with increased overlap (β = 0.22, p = 0.03), while low-lying peroneus brevis muscle showed no significant effect.

**Conclusion:** Peroneus brevis position is highly variable and depends on its shape, width, and the presence of peroneus quartus.

## Background

Ankle injuries are common and often complex, involving ligaments, cartilage, bone, or tendons injury. A frequent clinical challenge is the persistence of symptoms after ankle sprain, which can be caused by underdiagnosed peroneal tendon injuries (Michels et al., 2022; Wang et al., 2005). Peroneal tendon instability and split tears are both clinically and radiologically significant, yet they are frequently missed, with estimates suggesting that up to half of the peroneus brevis split tears go undiagnosed (Bokwa-Dabrowska et al., 2024; Davda et al., 2017). The true incidence of peroneus brevis instability is unknown. Some studies report a prevalence of peroneus brevis split tears as 30%–60% (Miura et al., 2004; Sobel et al., 1990). Textbooks typically describe the peroneus brevis at the ankle level as lying directly against the posterior surface of the lateral malleolus, with the peroneus longus coursing posterior to it. In clinical practice, however, this relationship is not always so straightforward.

Anatomical studies have investigated in detail the variations in the distal insertion of the peroneus brevis tendon and which anatomical variants may contribute to the development of peroneus brevis split tears. The distal attachment of the fibularis brevis tendon is known to demonstrate considerable morphological variability (Olewnik et al., 2020; Olewnik et al., 2024; Ruzik et al., 2022). The location of the musculotendinous junction and the risk of tendon injury. A cadaveric study found that a low-lying peroneus brevis muscle belly was statistically associated with longitudinal split tears of the fibularis brevis (Housley et al., 2017). Other anatomical variations related to peroneus brevis tears include the shape and depth of the fibular groove, presence of accessory muscles like the peroneus quartus, a low-lying muscle belly of the peroneus brevis, and hypertrophy of the peroneal tubercle (Galli et al., 2015; Mirmiran et al., 2015; Sobel et al., 1990; Sobel et al., 1993; Unlu et al., 2010; van Dijk et al., 2018; Wang et al., 2005).

Peroneal instability may occur with or without a tear of the superior peroneal retinaculum, which spans the posterior aspect of the lateral malleolus(Febriana and Aryana, 2023). Existing classifications of peroneal instability are based on imaging findings from transverse sections at this level(Oden, 1987). For this reason, we selected the level of the lateral malleolus as the reference point for our study. While peroneal tendon instability can be evaluated with ultrasound(Pesquer et al., 2016), its accuracy is dependent on the operator’s experience(Toprak et al., 2020). With the increasing use of magnetic resonance imaging and the development of dynamic MRI techniques(Garetier et al., 2020), there is a growing need for MRI-based studies. MRI provides a consistent and detailed assessment of peroneal tendon anatomy and plays an increasingly important role in ankle imaging. Although cadaveric studies have described distal insertional variants of the peroneus brevis tendon, no in vivo MRI-based study has systematically evaluated its position relative to the peroneus longus at the level of the superior peroneal retinaculum, where instability commonly arises.

Our study addresses this gap by investigating the anatomical variants in the position of the peroneus brevis tendon relative to the peroneus longus in vivo on magnetic resonance imaging at the level of the lateral malleolus. By analysing a cohort of asymptomatic individuals using high-resolution imaging, we aim to improve the understanding of normal anatomical variability and its potential relevance to peroneal tendon instability.

## Objectives

To classify anatomical variants in the position of the peroneus brevis tendon relative to the peroneus longus at the level of the lateral malleolus, and to investigate associations with tendon shape, cross-sectional area, and the presence of peroneus quartus or a low-lying muscle belly.

*Research questions:*

1. What are the most common anatomical variants in the relative position of the peroneus brevis tendon to the peroneus longus?
2. Are shape, cross-sectional area, the presence of peroneus quartus, and low-lying muscle belly related to the position of peroneus brevis?

## Methods

### 1. Study design

This is an observational cohort study. The study was conducted and reported according to the Strengthening the Reporting of Observational Studies in Epidemiology (STROBE) guidelines for observational studies (von Elm et al., 2007).

### 2. Setting

We re-evaluated ankle MRI examinations performed on a 3T scanner between November 2017 and December 2024 at our institution using standard protocol.

### 3. Participants

Inclusion criteria were normal peroneal tendons and normal superior peroneal retinaculum. Exclusion criteria included higher signal of peroneus brevis or longus on proton density weighted or T2-weighted images, tenosynovitis in the peroneal synovial sheath and bone marrow oedema, lateral malleolar fracture and symptoms in relation to lateral malleolus in the referral letter, history of peroneus tendon pathology, prior surgery, inflammatory conditions, or tumours.

Participants were excluded if MRI demonstrated peroneal tendon pathology, defined as abnormal signal on T2-or proton density–weighted images suggestive of a split tear, tenosynovitis, or tendinosis. Additional exclusion criteria included fractures, local infections, ganglion cysts or tumors, transverse rupture of the peroneus brevis, rupture of the peroneus longus, postoperative changes, metal or MRI-related artifacts, duplicate MRI examinations from the same individual, missing anthropometric data, and age under 18 years.

### 4. Outcomes

**The primary outcome** is the relative position of the peroneus brevis tendon in relation to the peroneus longus tendon at the level of the lateral malleolus, assessed on axial magnetic resonance imaging. Position was categorised as:

a) peroneus brevis overlaps the peroneus longus and protrudes medially;
b) peroneus brevis is positioned entirely medial to the peroneus longus;
c) peroneus brevis overlaps the peroneus longus and protrudes laterally; and
c) peroneus brevis overlaps the peroneus longus and protrudes both medially and laterally.

**Secondary outcomes** are shape, cross-sectional area of the peroneus brevis tendon and presence of the peroneus quartus muscle.

The shape of the peroneus brevis tendon classified into four types based on cross-sectional morphology:

a) general flat
b) flattened with lateral convexity
c) flattened with medial convexity
c) oval

Cross-sectional area of the peroneus brevis tendon (in mm²), measured on axial images.

**Potential confounders**: age, sex, and side of the body (left or right ankle).

### 5. Data sources and measurements

MRI transverse sections (proton density-weighted) 1 cm above the lateral malleolus apex were identified by two independent musculoskeletal radiologists. Discrepancies were resolved by consensus. The sections were segmented using validated macro in ImageJ (Appendix A), where the outlines of the peroneus brevis, peroneus longus, and fibula were segmented by two raters (musculoskeletal radiologist with 10 years’ experience and physiotherapist after getting a training session from musculoskeletal radiologist) to obtain coordinates (X, Y) allowing to assess the cross-sectional areas of the tendons (mm2) and overlap (mm).

### Rater training and calibration

Prior to data collection, the two raters (a musculoskeletal radiologist with 10 years of experience and a physiotherapist trained in musculoskeletal imaging) completed a joint calibration session using a sample set of 10 MRI examinations not included in the study. During this session, the raters reviewed image quality, anatomical boundaries, segmentation procedures, and measurement protocols.

### 6. Sample size

A sample size of 230 MRI examinations was selected based on our preliminary study on 50 examinations to achieve the power for the chi-square test (as the main analysis) of more than 0.8. The effect size, measured using Cramér’s V, was 0.38.

### 7. Quantitative analysis

Tendon area (in mm²) as a continuous variable was compared between four tendon shapes using analysis of variance (ANOVA).

### Measurement of tendon width

Tendon width was defined as the maximum mediolateral distance across the peroneus brevis tendon on the selected transverse section. Measurements were taken orthogonal to the long axis of the tendon, based on manually segmented outlines in ImageJ. The width was automatically calculated from the segmented region using the validated macro (Appendix A).

### 8. Statistical analysis

Inter-rater agreement was assessed using Cohen’s kappa for categorical variables (localization and shape), and the intraclass correlation coefficient (ICC(2,1)) for continuous measurements (cross-sectional area).

Chi-square tests were used to assess the association between the relative position of the peroneus brevis tendon and its shape, as well as between age groups (18–30 years, 31–60 years, and older than 61 years) and peroneus brevis shape. In case of a significant chi-square result, Tukey’s HSD post hoc test was applied to identify specific group differences. Effect sizes for chi-square tests were reported using Cramér’s V with 95% confidence intervals.

ANOVA was used to evaluate if cross-sectional area of the peroneus brevis tendon differed between different shape and positions. The assumption of normality was evaluated using the Shapiro–Wilk test, and homogeneity of variance was assessed with Levene’s test. A significance level of p < 0.05 was considered significant. Post-hoc comparisons were conducted using Tukey’s honest significant difference (HSD) test. Effect sizes for ANOVA were calculated using eta squared (η²) with 95% confidence intervals.

The distribution of overlap values was non-normal in at least two groups (Shapiro–Wilk p < 0.001) and Levene’s test indicated significant heterogeneity of variances (p = 0.0037). Therefore, we used the Kruskal–Wallis test followed by pairwise Wilcoxon tests with Benjamini–Hochberg correction. Where appropriate, non-parametric effect sizes (e.g., epsilon squared or rank-based correlations) were also reported.

We used a multiple linear regression model to assess association between variables and the degree of tendon overlap (mm). Predictor variables included in the regression model were selected based on anatomical plausibility and evidence prior studies suggesting a potential association with tendon pathology. No variable selection procedures (e.g., stepwise selection) were applied post hoc; all predictors were pre-specified. The regression model was constructed using four predictors: presence of a peroneus quartus muscle, presence of a low-lying peroneus brevis muscle belly, cross-sectional area of the peroneus longus tendon, and width of the peroneus brevis tendon. Given evidence of non-constant variance (Breusch– Pagan test, p < 0.001), robust standard errors (HC1 estimator) were applied to all coefficient estimates. Model assumptions were evaluated through residual plots and normality testing (Shapiro–Wilk, p = 0.026). Model robustness was assessed by influence diagnostics and sensitivity analysis excluding the most influential observation (one case).

Model assumptions were assessed using residual, Q–Q, and scale–location plots. Residuals showed approximate normality and homoscedasticity. Cook’s distance identified moderately influential cases (threshold 4/n), which were reviewed but retained, as sensitivity analysis showed no impact on conclusions. We assessed multicollinearity between predictors using Pearson correlation coefficients and variance inflation factors (VIF). A correlation matrix was generated (Supplementary Table S4), and VIF values were calculated for all predictors in the final regression model.

Statistical analyses were conducted using R (version 4.3.2; R Foundation for Statistical Computing, Vienna, Austria) with the following packages: stats, car, sandwich, lmtest, rstatix, irr, psych, effectsize, rcompanion, ggplot2, and dplyr.

## Results

### 1. Study population

We re-evaluated 749 MRI examinations and 230 consecutive MRI examinations met the inclusion criteria, Figure 1.

**Figure 1.**
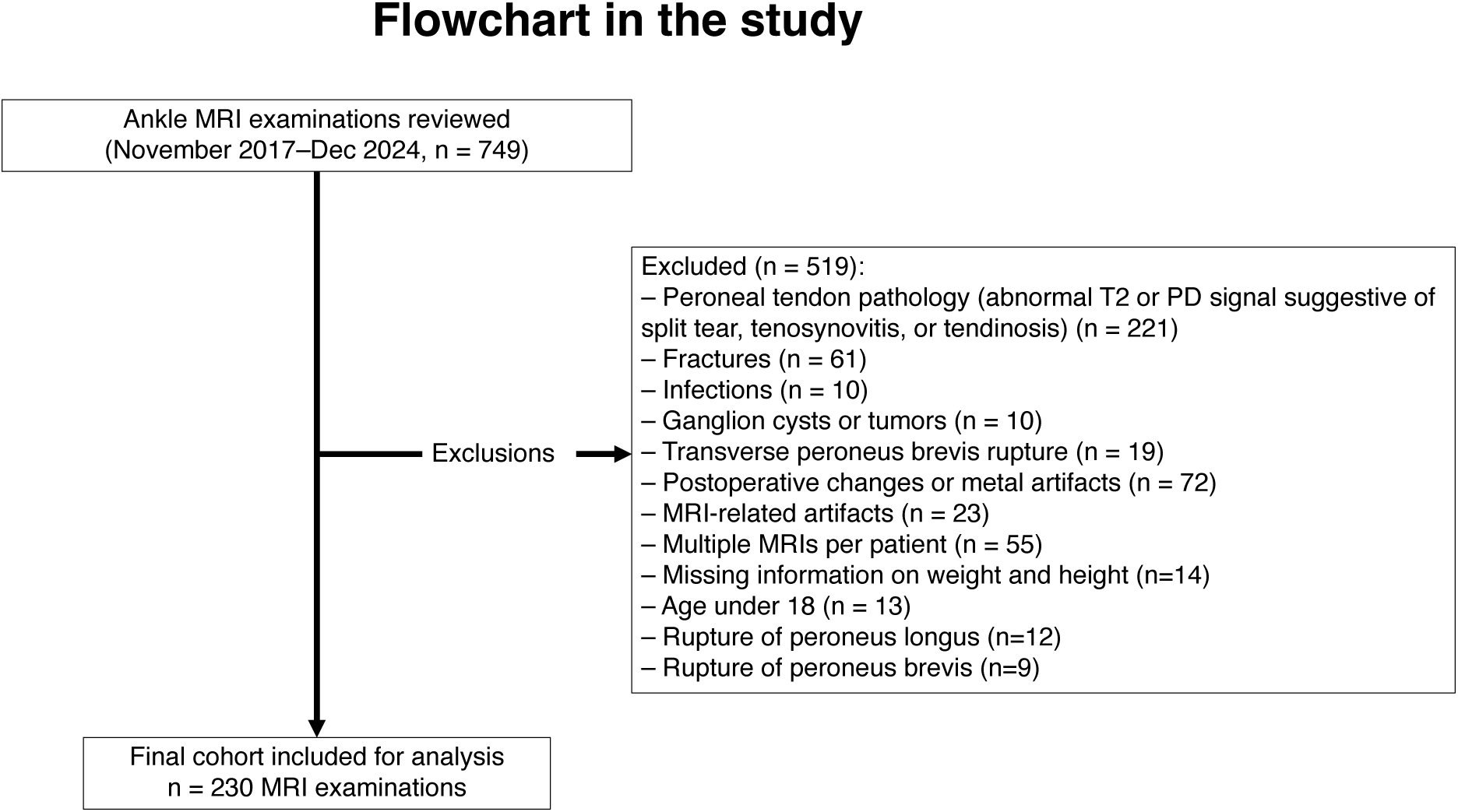
Flowchart.

The mean age was 41.9 years (range: 12–72 years), with a standard deviation of 14.9 years. Among the total of 230 patients, there were 121 males (52.6%) and 109 females (47.4%). The distribution of side was equal, with 115 patients on the left side (50.0%) and 115 patients on the right side (50.0%).

### 2. Outcome data and main results

#### 2.1. Inter-rater agreement

Inter-rater agreement for the localization, shape, and cross-sectional area of the peroneus brevis tendon was almost perfect, according to the criteria by Landis and Koch (Landis and Koch, 1977). Cohen’s kappa was 0.85 (95% CI: 0.76–0.94) for localization and 0.87 (95% CI: 0.81–0.98) for shape. The intraclass correlation coefficient (ICC(2,1)) for cross-sectional area was 0.98 (95% CI: 0.97–0.99). Agreement for identifying a low-lying muscle belly (LLMB) was also almost perfect (kappa = 0.83, 95% CI: 0.73–0.92), while agreement for the presence of a peroneus quartus was moderate (kappa = 0.66, 95% CI: 0.54–0.78) (Landis and Koch, 1977). Final ratings for localization, shape, LLMB, and peroneus quartus were based on consensus; for cross-sectional area, the mean value was used due to high agreement.

#### 2.2. Variants in the position of the peroneus brevis tendon

The most common relative position was peroneus brevis overlaps peroneus longus and protrudes medially, observed in 161 cases (72.0%). The second most frequent position was peroneus brevis medial to peroneus longus, seen in 33 cases (15.1%), followed by peroneus brevis overlaps peroneus longus and protrudes laterally, observed in 22 cases (9.6%). The least common position was peroneus brevis overlaps peroneus longus and protrudes both laterally and medially, noted in 14 cases (6.0%) (Figure 2). The peroneus brevis tendon position was not significantly influenced by age, sex, or side (p > 0.05).

**Figure 2.**
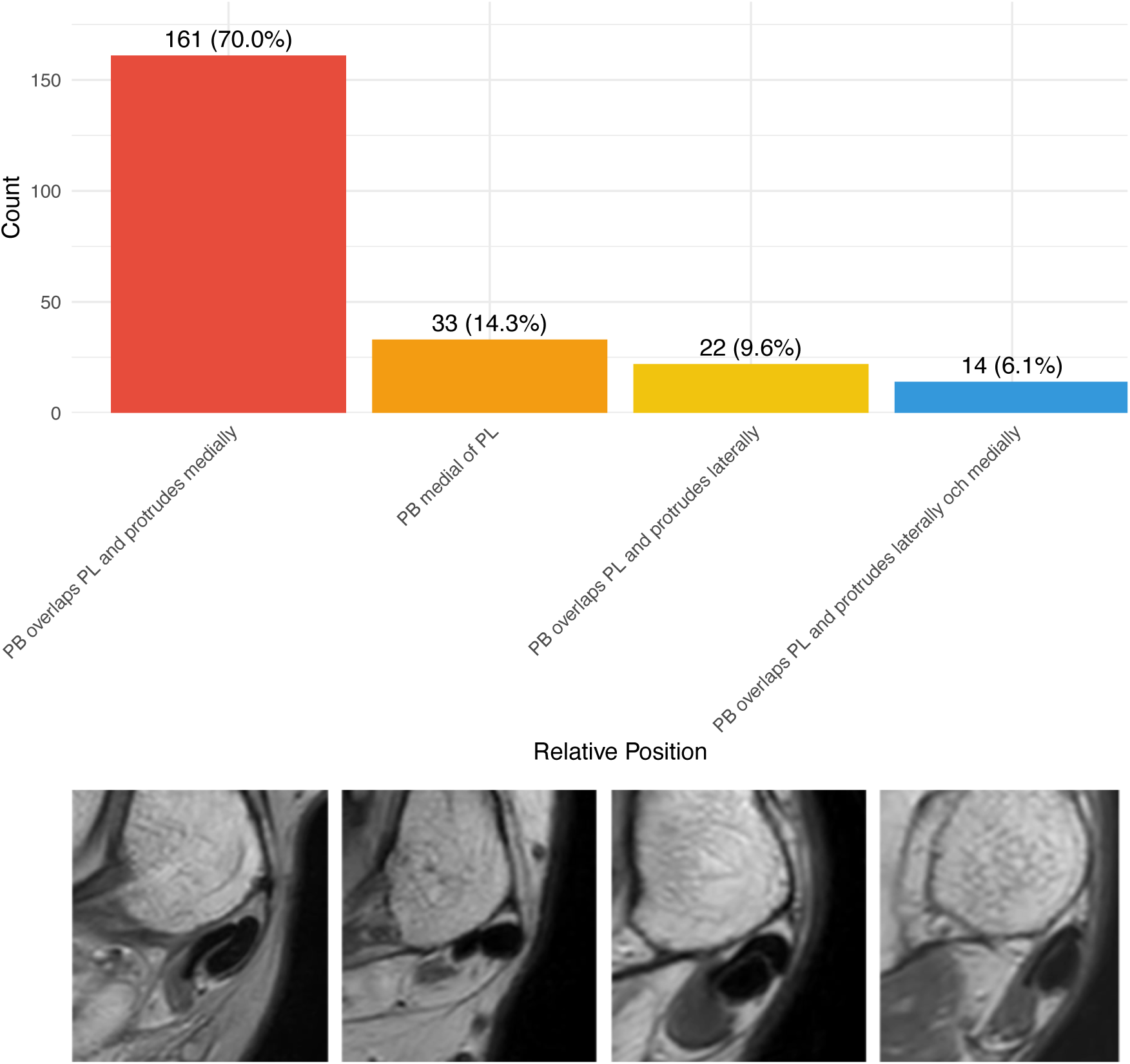
Bar chart illustrating the distribution of relative tendon positions with counts (n) and percentages (%). Examples from the magnetic resonance imaging (MRI), proton density-weighted images, transverse sections showing peroneal tendons in relation to lateral malleolus, left ankles. MRI sections below correspond to the bar chart above, with the same order and legend for tendon position. PB – peroneus brevis, PL – peroneus longus.

#### 2.3. Tendon shape vs. position

The most frequent shape across all positions was general flat (82 cases, 36.3%), followed by flattened convex medially (81 cases, 35.8%), oval (42 cases, 18.3%), and flattened convex laterally (25 cases, 9.6%) (Figures 3 and 4).

**Figure 3.**
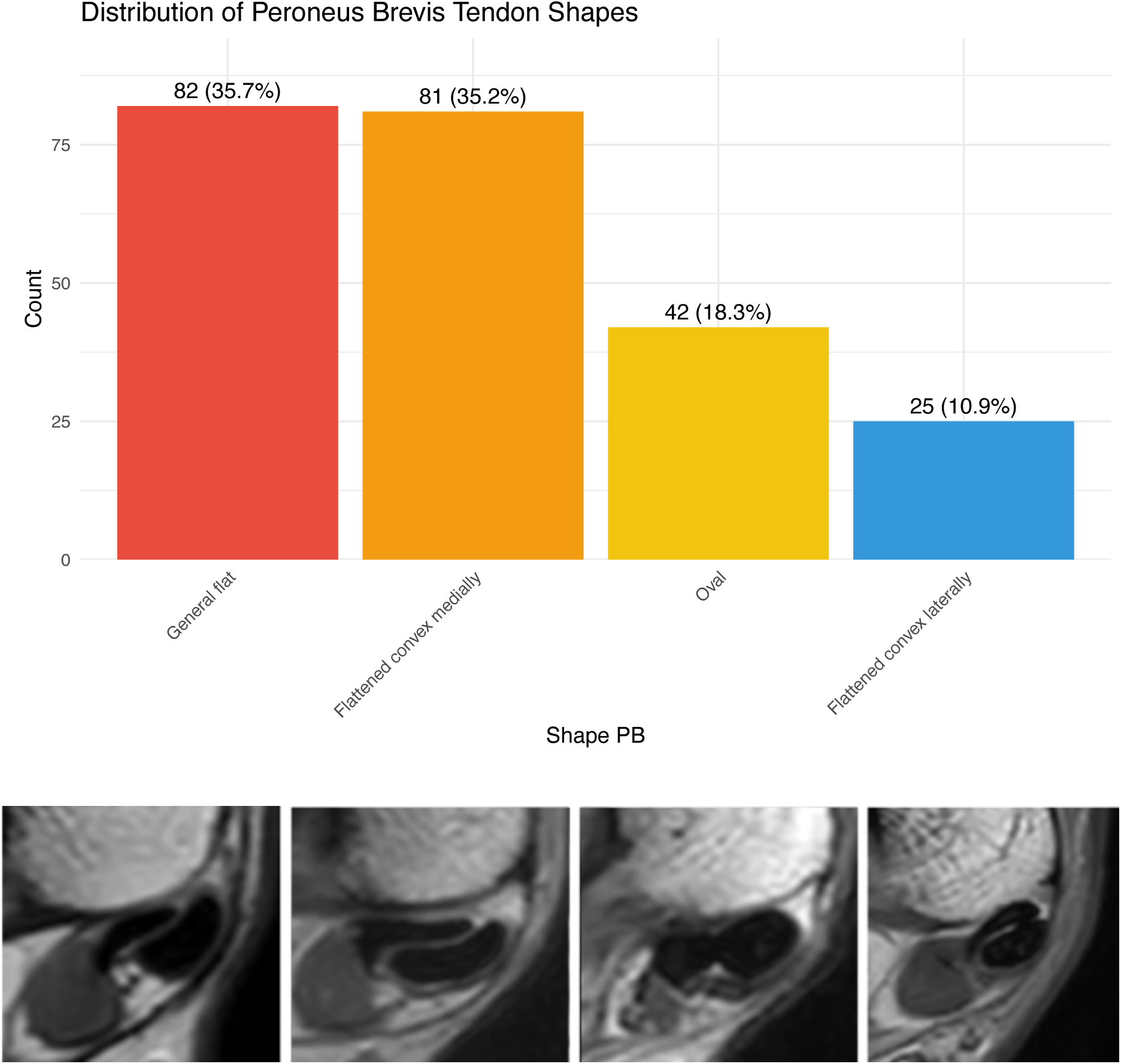
Bar chart showing the distribution of tendon shapes with examples from proton density weighted images. Examples from the magnetic resonance imaging (MRI), proton density-weighted images, transverse sections showing peroneal tendons in relation to lateral malleolus, left ankles. MRI sections below correspond to the bar chart above, with the same order and legend for tendon position. PB – peroneus brevis.

**Figure 4.**
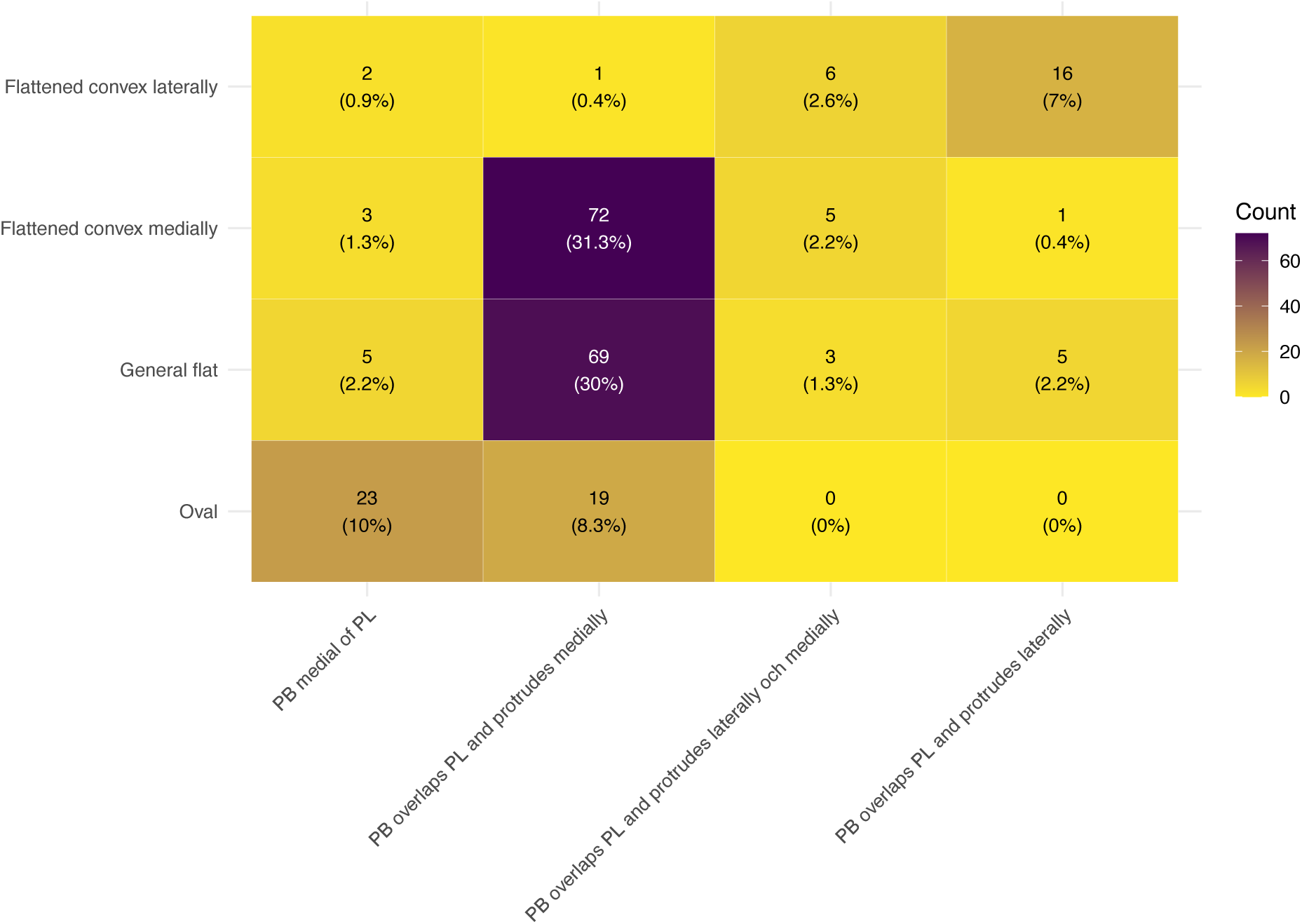
Distribution of peroneus brevis shape by position, transition matrix.

There was a significant association between the relative position of the peroneus brevis tendon and its cross-sectional shape (chi2 χ² = 189.41, df = 9, p < 0.001). Peroneus brevis located medial to the peroneus longus were most commonly oval-shaped, while those overlapping and protruding medially were frequently flattened convex medially or general flat (Figures 4 and 5). These findings suggest a morphological relationship between peroneus brevis shape and anatomical position.

**Figure 5.**
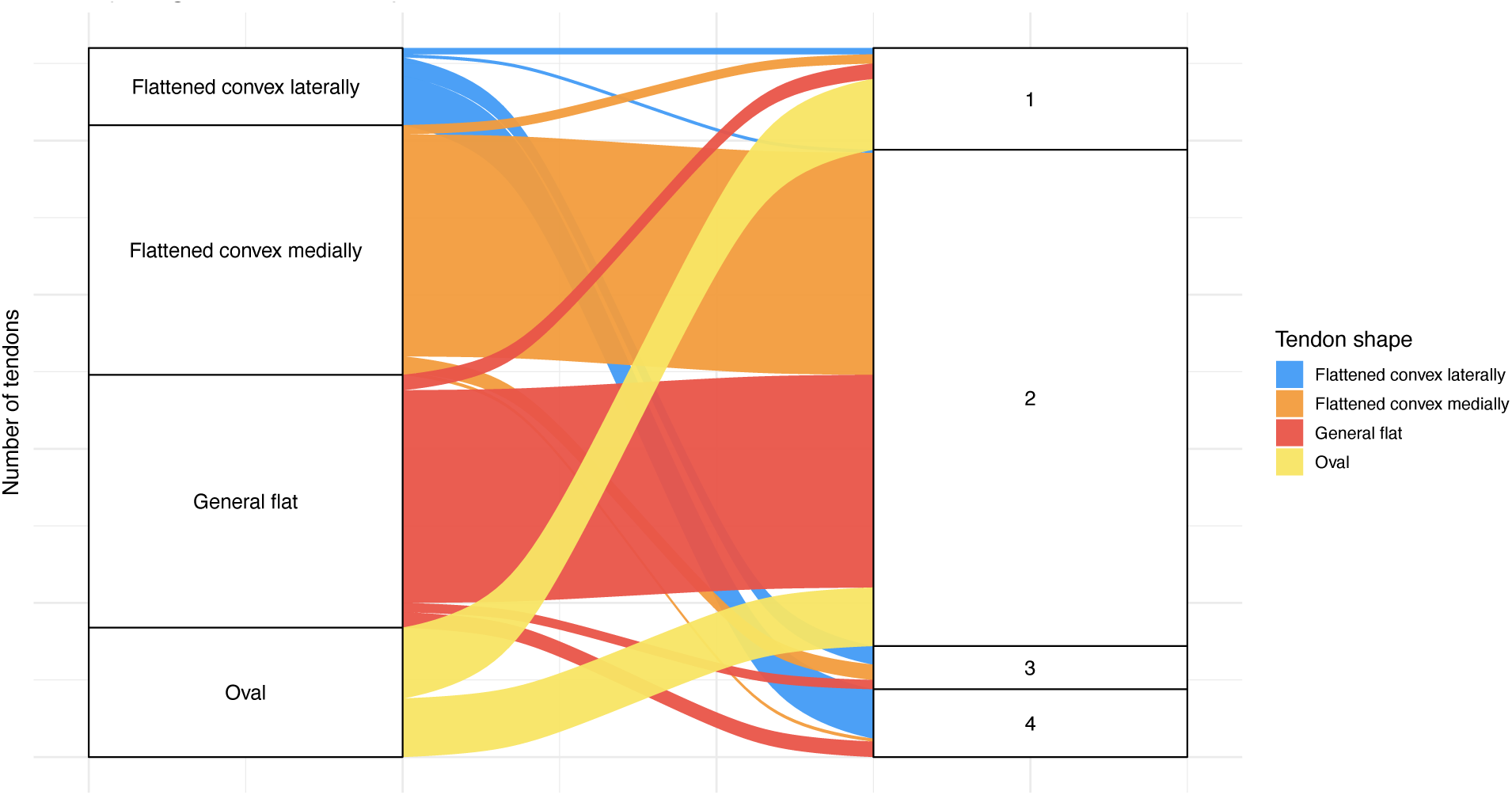
Sankey diagram illustrating the distribution of tendon shapes in relation to their position (1–4). The width of each flow is proportional to the number of tendons in each combination. On the left, the shape of the peroneus brevis tendon is shown, categorised as flattened convex laterally (blue), flattened convex medially (orange), general flat (red), or oval (yellow). These shapes are mapped to the tendon’s relative position to the peroneus longus on the right side of the plot. 1 – peroneus brevis medial to peroneus longus, 2 – peroneus brevis overlaps peroneus longus and protrudes medially, 3 – peroneus brevis overlaps peroneus longus and protrudes both laterally and medially, and 4 – peroneus brevis overlaps peroneus longus and protrudes laterally

**Figure 6.**
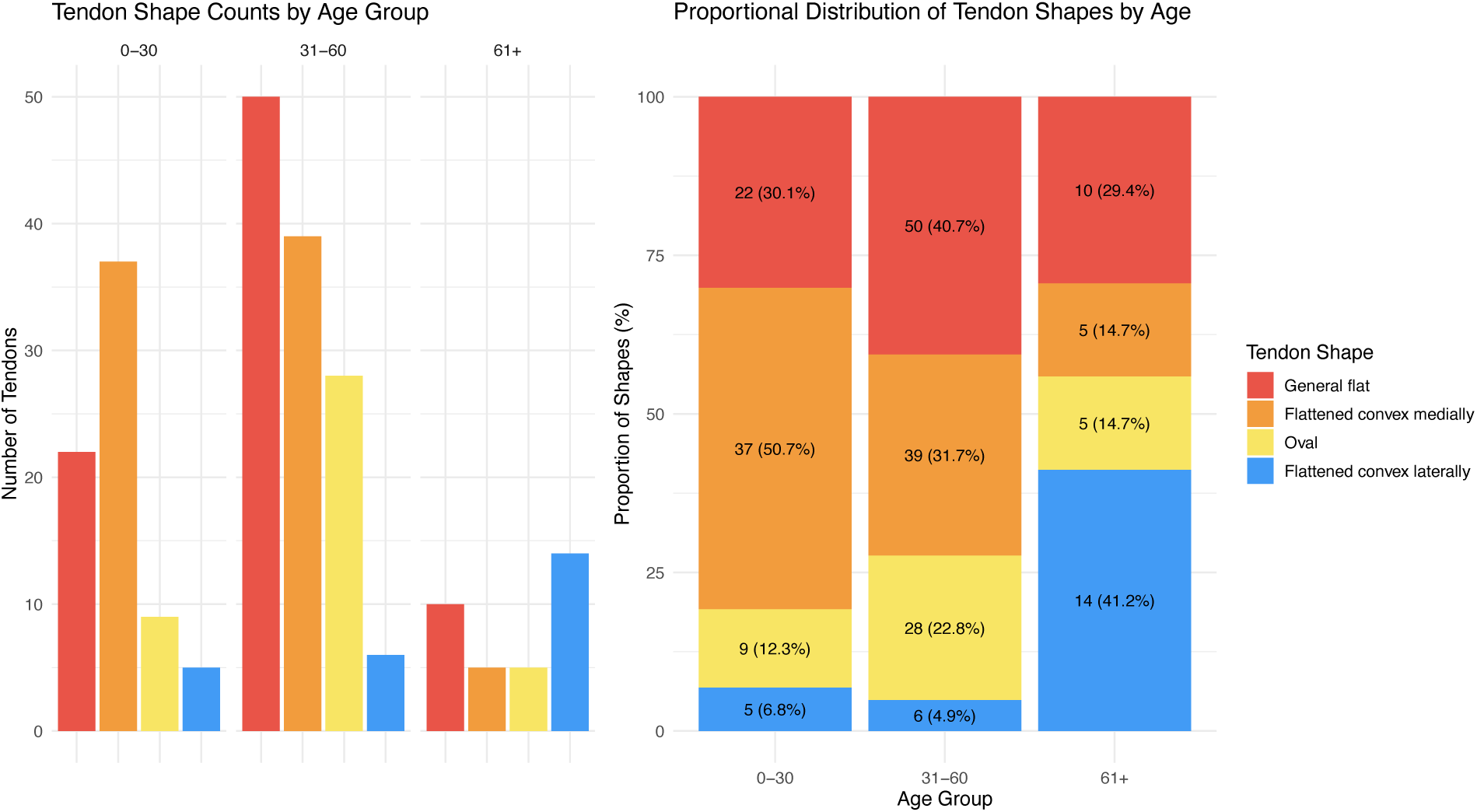
Combined visualisation of peroneus brevis tendon shape distribution across age groups. (Left) Absolute counts of each shape within age groups. (Right) Proportional distribution, highlighting shape frequency normalised to group size. Colour-coding corresponds to tendon shape: general flat (red), flattened convex medially (orange), oval (yellow), and flattened convex laterally (blue).

#### 2.4. Tendon shape vs age group

The ‘flattened convex medially’ shape was most common in individuals aged 31–60 years (31.7%), while the ‘flattened convex laterally’ shape appeared more frequently in those over 61 years (38.9%). A Chi-square test confirmed a significant association between age group and peroneus brevis shape (χ² = 11.03, df = 9, p = 0.004). There was a significant association between peroneus brevis tendon shape and age group (χ² = 48.19, df = 6, p < 0.001, Cramér’s V = 0.32, 95% CI: 0.22–0.45), indicating a moderate effect size. Post-hoc analysis (Tukey’s HSD) indicated that this difference was driven by comparisons between the 31–60 and 61+ age groups (mean difference = 2.24, p = 0.036) (Figure 7).

**Figure 7.**
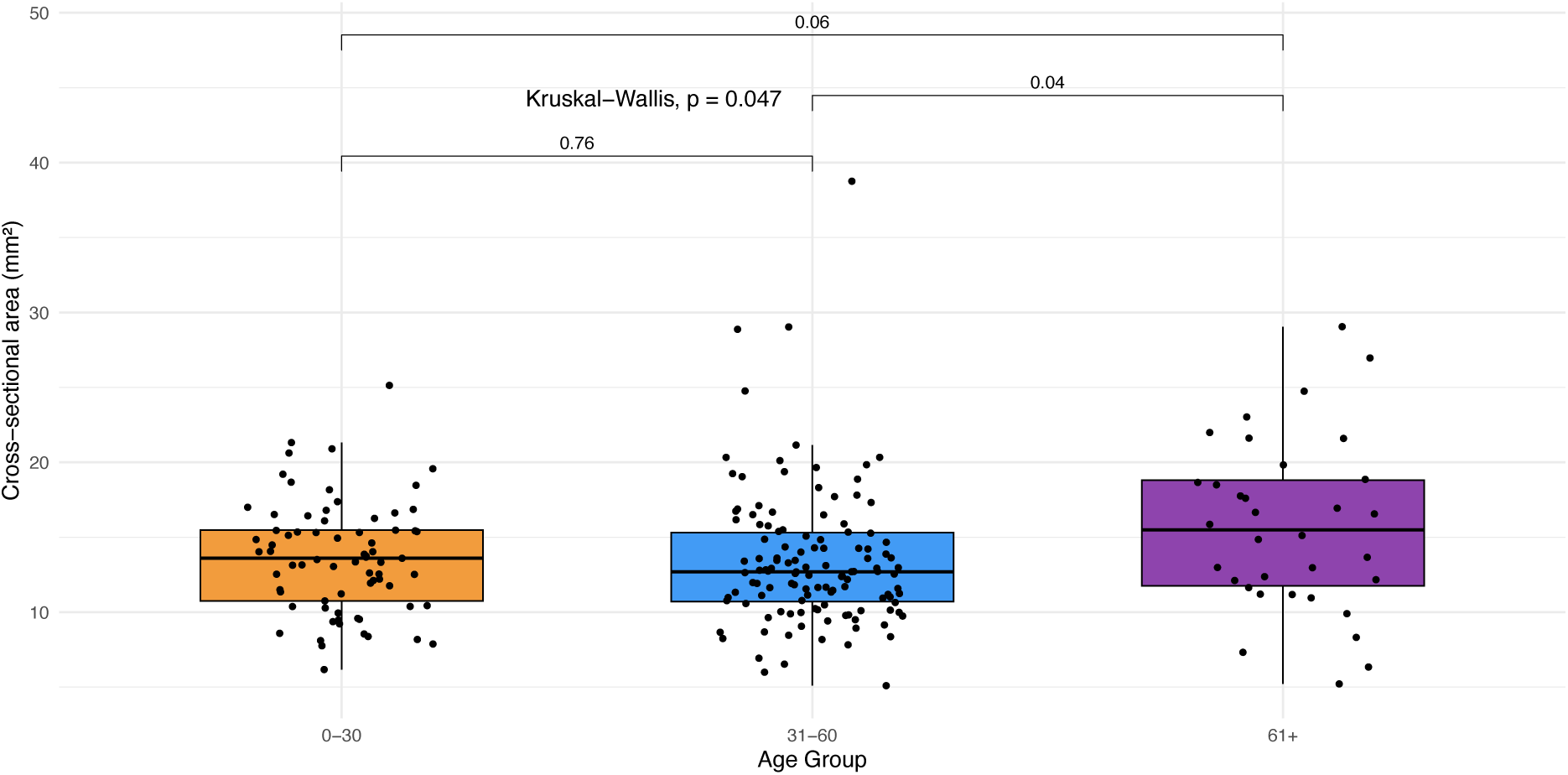
Peroneus brevis cross sectional area by age group. Boxplot showing the distribution of peroneus brevis tendon cross-sectional area (mm²) across age groups. Coloured boxes represent age categories: 0–30 (orange), 31–60 (blue), and 61+ (purple). Black dots indicate individual measurements. A Kruskal–Wallis test revealed a significant overall difference between groups, with pairwise comparisons indicating a significant difference between individuals aged 31–60 and those over 61 (p = 0.036).

#### 2.5. Relation between tendon position and cross-sectional area

The mean peroneus brevis cross-sectional area (mm²) varied across different tendon positions (Table 1). The highest cross section area of the peroneus brevis was seen when it overlaps peroneus longus and protrudes laterally and medially (mean area of 16.9 mm², SD = 5.7).

**Table 1.** Differences in the cross-sectional area of peroneus brevis vs different positions of the peroneus brevis tendon.

| Relative Position | Count | Mean | SD | Min | Q1 | Median | Q3 | Max |
| --- | --- | --- | --- | --- | --- | --- | --- | --- |
| PB medial of PL | 33 | 12.2 | 3.7 | 5.1 | 10.2 | 11.2 | 13.5 | 25.1 |
| PB overlaps PL and protrudes laterally | 22 | 14.8 | 5.3 | 6.3 | 10.4 | 15 | 18.1 | 27 |
| PB overlaps PL and protrudes laterally and medially | 14 | 16.9 | 5.7 | 9.7 | 12.3 | 15.7 | 20.3 | 29 |
| PB overlaps PL and protrudes medially | 161 | 13.8 | 4.4 | 5.2 | 11.2 | 13.1 | 15.9 | 38.8 |

The cross-sectional area of the peroneus brevis tendon differed significantly between position groups (ANOVA, F(3, 226) = 3.96, p = 0.0088, η² = 0.05, 95% CI: 0.01–1.00), indicating a small effect size. Post-hoc pairwise comparisons (Bonferroni-adjusted t-tests) showed a significantly larger area in peroneus brevis overlapping and protruding medially compared to those located medial to the peroneus longus (p = 0.032; Figure 8). No other pairwise differences reached statistical significance.

**Figure 8.**
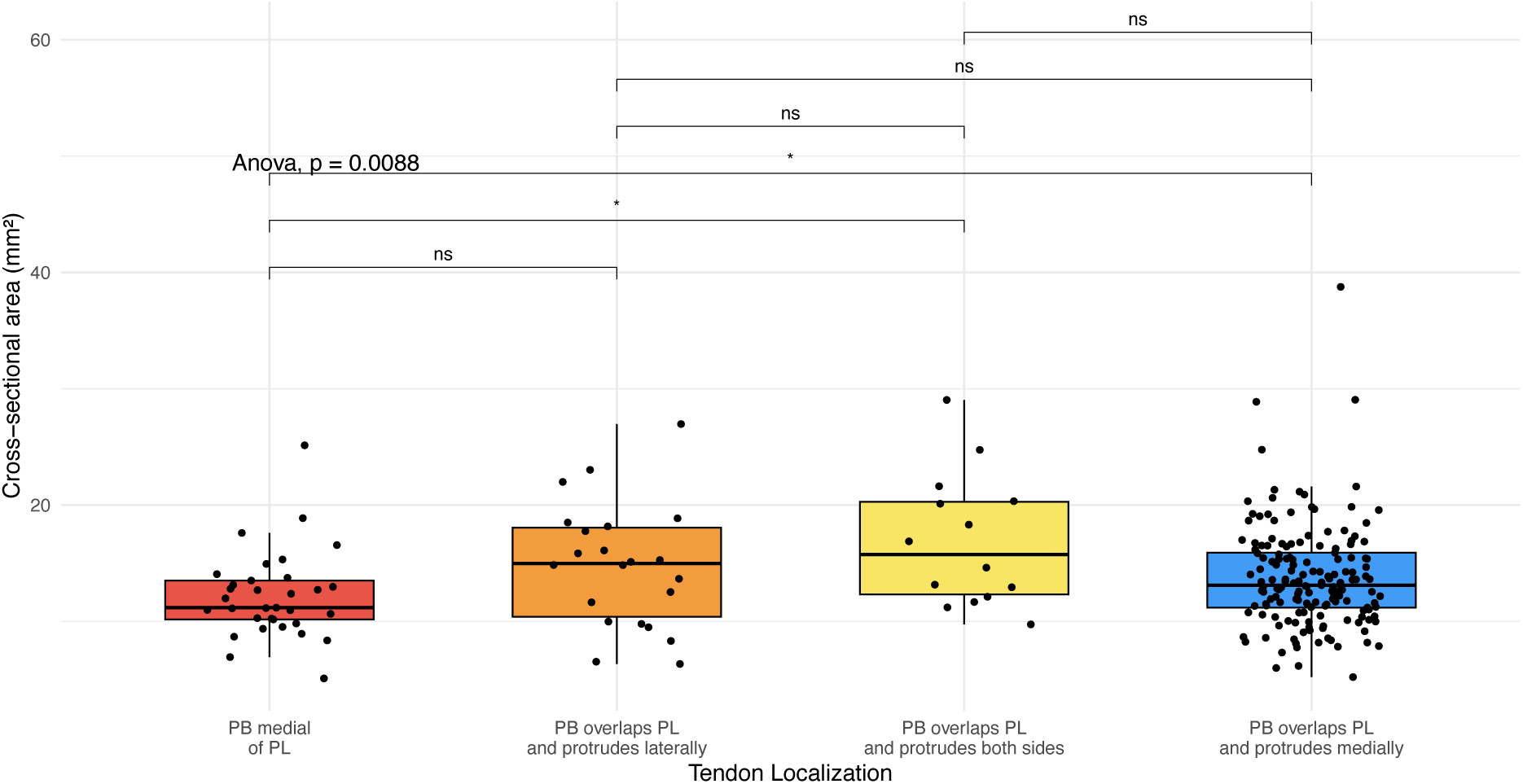
Peroneus brevis cross section area by tendon position, boxplot showing. Coloured boxes indicate position categories, and individual data points are overlaid with jitter. An overall group difference was detected using one-way analysis of variance (ANOVA) (p = 0.0088). Pairwise comparisons were assessed using Bonferroni-adjusted t-tests. Significant differences are marked with asterisks; non-significant comparisons are labelled ‘ns’. PB – peroneus brevis, PL – peroneus longus.

#### 2.6. The relationship between tendon shape and cross-sectional area

The highest mean cross-sectional area of the peroneus brevis tendon was observed in tendons with a flattened convex lateral shape (mean ± SD: 16.5 ± 6.3 mm²; Table 2).

**Table 2.**
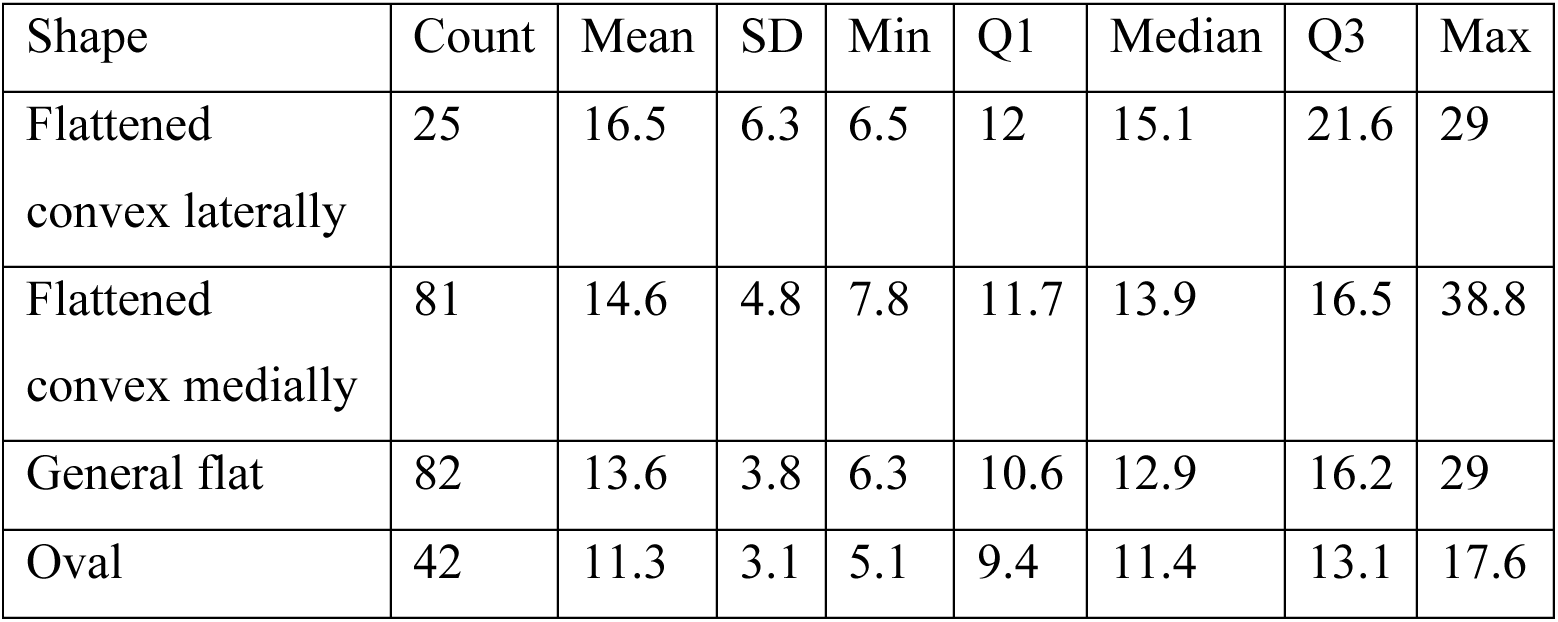
The relationship between peroneus brevis shape and cross-sectional area (in mm^2^).

A one-way ANOVA revealed a statistically significant differences in cross-sectional area between tendon shape categories (F(3, 226) = 8.73, p < 0.001). A post-hoc Tukey’s test showed that oval-shaped tendons had a significantly smallest cross-sectional area (Figure 9).

**Figure 9.**
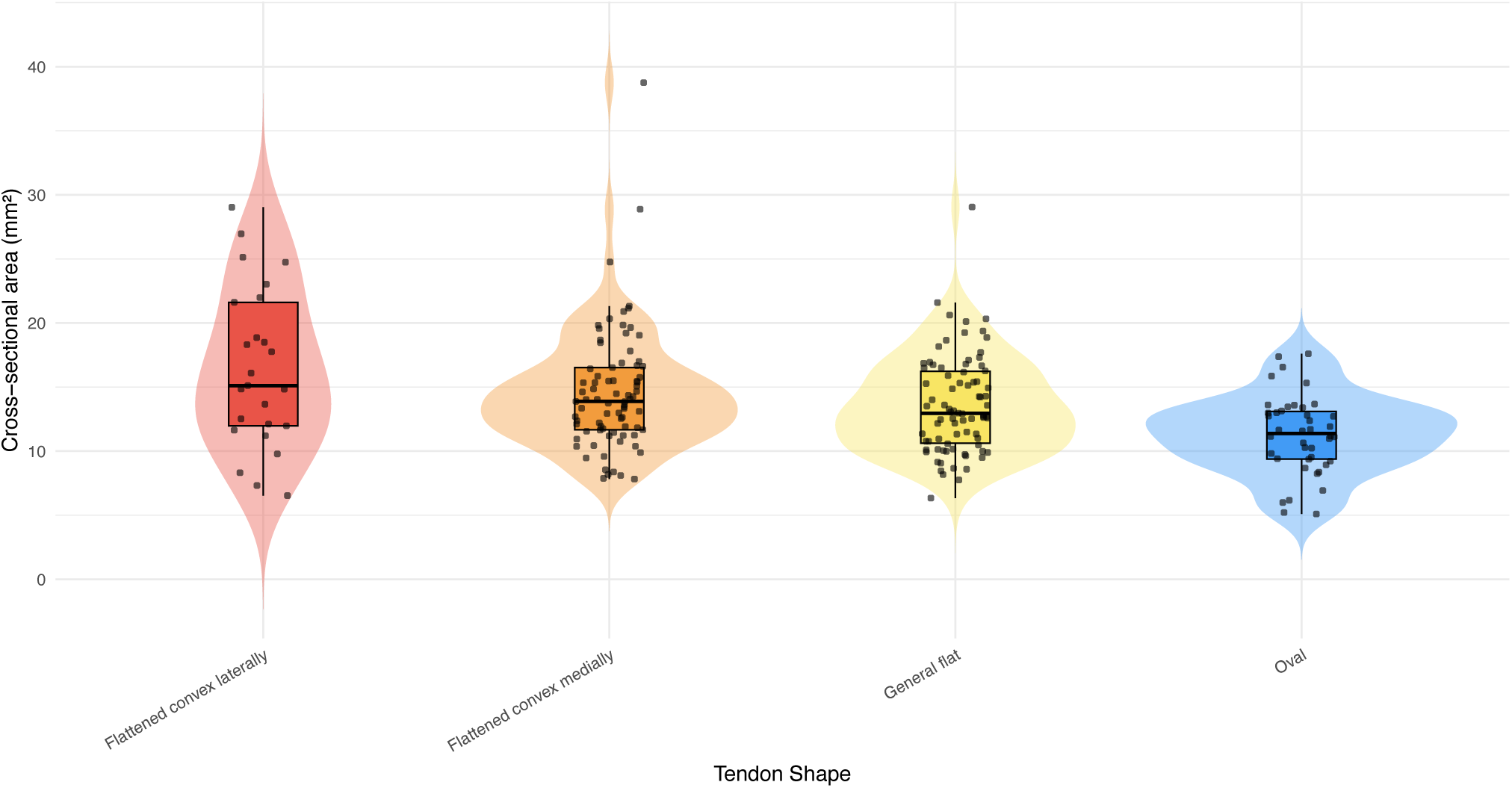
Violin plot showing the distribution of peroneus brevis tendon cross-sectional area across different tendon shapes. Boxplots indicate median and interquartile range. Jittered dots represent individual tendons.

#### 2.7. Grade of the overlap between tendons

The degree of overlap between the peroneus brevis and longus tendons differed significantly across the relative position groups of the peroneus brevis, as shown by Fisher’s exact test (p < 0.001), one-way ANOVA (F(3, 226) = 46.96, p < 0.001), and the Kruskal–Wallis test (χ² = 91.2, p < 0.001).

The greatest overlap occurred where the peroneus brevis overlapped the longus and protruded laterally (mean ± SD: 3.2 ± 0.9 mm) and both laterally and medially (3.1 ± 0.7 mm). Almost no overlap was observed when the peroneus brevis was located medial to the longus (0.6 ± 0.6 mm; Table 3). Post hoc analysis (Tukey HSD) showed statistically significant differences between all groups, except between the two configurations with the greatest overlap (Table 3, Figure 10).

**Figure 10.**
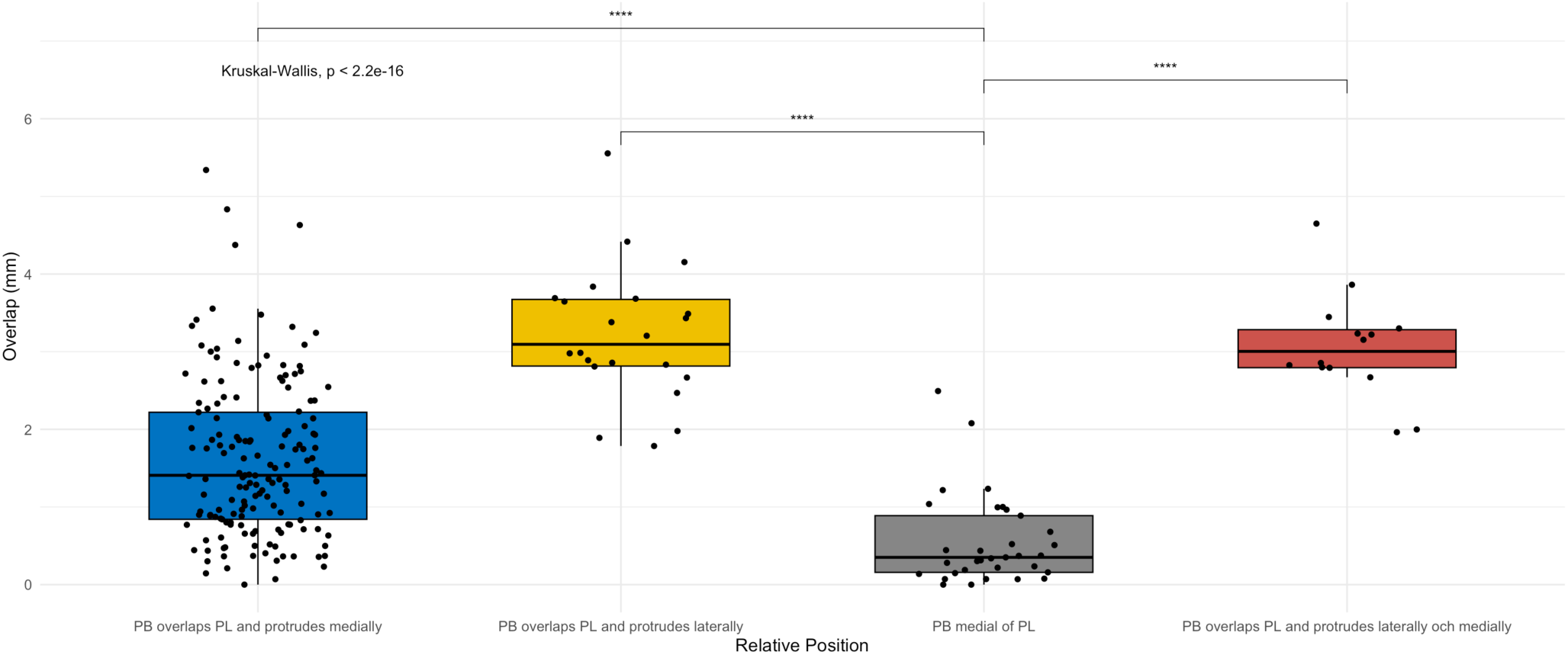
Overlap in mm across tendon positions. Boxplots display the distribution of measured overlap between the peroneus brevis and peroneus longus tendons across four anatomical positions. A Kruskal–Wallis test showed a significant overall difference (p < 2.2×10⁻¹⁶), and post hoc pairwise Wilcoxon tests (Benjamini–Hochberg adjusted) confirmed significantly greater overlap in all overlapping variants compared to the medial position (**** = p < 0.0001). PB – peroneus brevis, PL – peroneus longus.

**Table 3.** Degree of overlap (in mm) between the peroneus brevis and longus tendons across different relative tendon positions. Values are presented as mean ± standard deviation (SD).

| Relative Position | Count | Mean | SD | Mean ± SD |
| --- | --- | --- | --- | --- |
| PB medial of PL | 33 | 0.6 | 0.6 | $0.6 \pm 0.6$ |
| PB overlaps PL and protrudes laterally | 22 | 3.2 | 0.9 | $3.2 \pm 0.9$ |
| PB overlaps PL and protrudes laterally and medially | 14 | 3.1 | 0.7 | $3.1 \pm 0.7$ |
| PB overlaps PL and protrudes medially | 161 | 1.6 | 1 | $1.6 \pm 1$ |

PB – peroneus brevis, PL – peroneus longus.

Greater tendon width (6.80 mm vs 5.48 mm, p < 0.001) was strongly associated with increased overlap between the peroneus tendons. Width showed the strongest correlation with overlap (r = 0.79), followed by area (r = 0.35), and thickness (r = 0.22) (all p < 0.001; Table 4, Figure 12).

**Figure 11.**
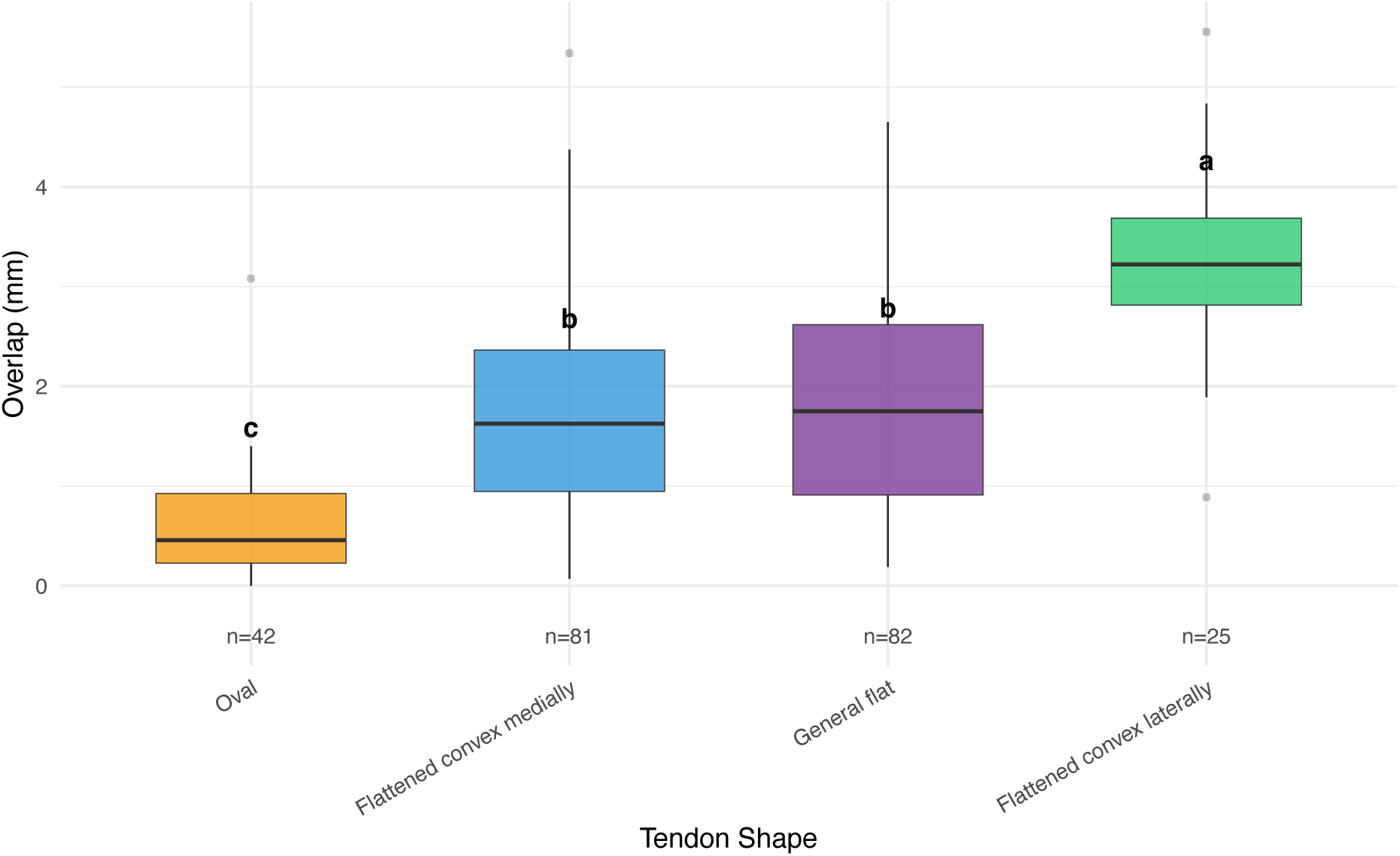
Overlap (in mm) between the peroneus brevis and longus tendons across different tendon shapes. Each boxplot shows the distribution of overlap values within each shape group, including the median, interquartile range, and outliers. Letters above the boxes indicate statistically significant groupings based on Tukey’s HSD post-hoc analysis (p < 0.05): Groups that share the same letter were not significantly different, while groups with different letters differed significantly. The sample size for each shape group is displayed below the x-axis.

**Figure 12.**
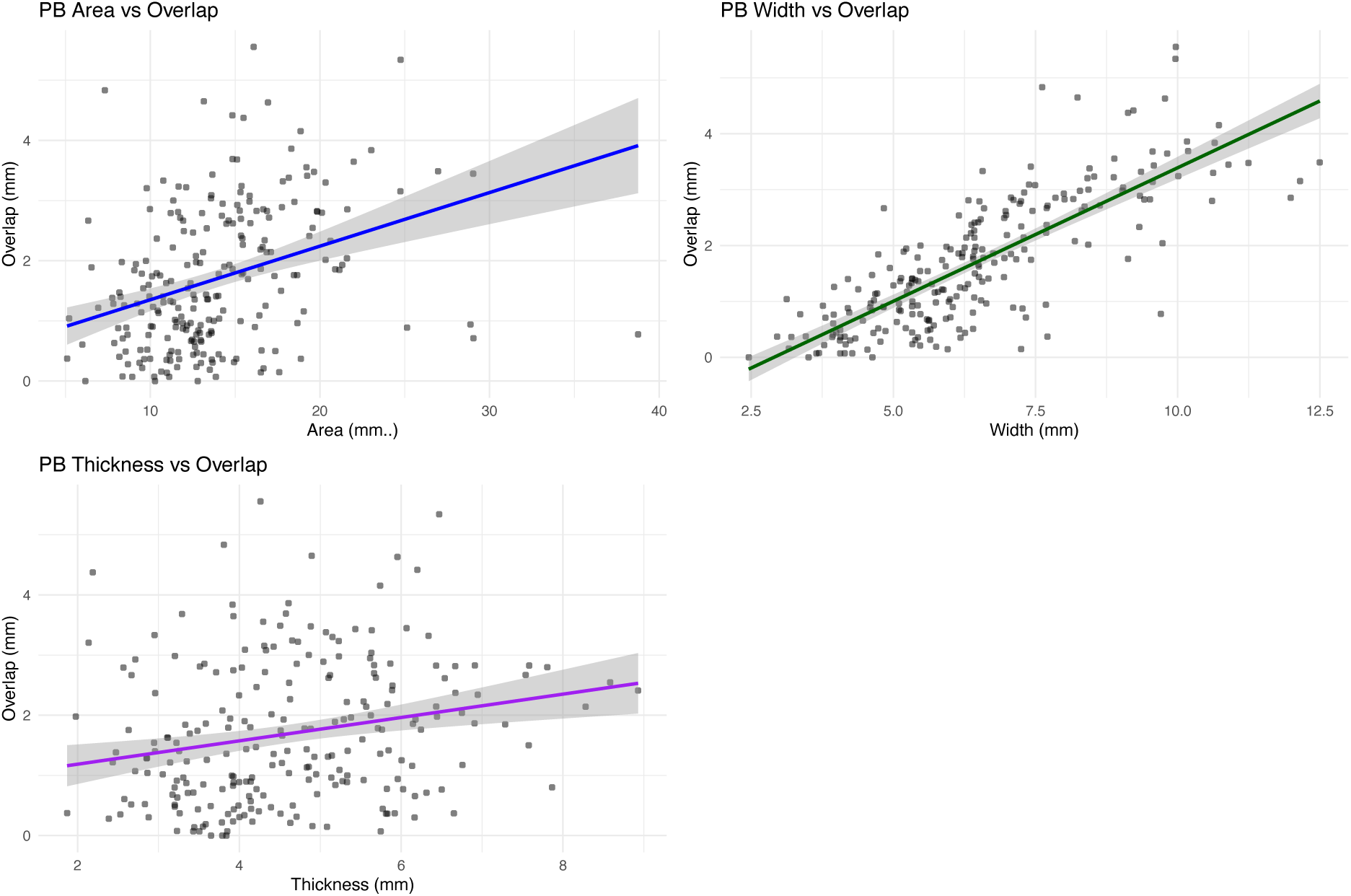
The relationship between the peroneus brevis tendon overlap and its cross-sectional area, thickness, and width. (A) Cross-sectional area, (B) width, and (C) thickness of the peroneus brevis tendon are plotted against the measured overlap in millimetres. Each dot represents a single patient, and the blue line indicates the linear regression fit with a 95% confidence interval (CI). Width showed the strongest correlation with overlap (r = 0.79), followed by area (r = 0.35) and thickness (r = 0.22). PB – peroneus brevis.

**Table 4.** Pearson correlation between peroneus brevis tendon dimensions (cross-sectional area, width, and thickness) and overlap with the peroneus longus tendon.

| Variable | Pearson (r) | 95% CI lower | 95% CI upper | p value |
| --- | --- | --- | --- | --- |
| PB Area vs Overlap | 0.35 | 0.23 | 0.46 | < 0.001 |
| PB Width vs Overlap | 0.79 | 0.74 | 0.83 | < 0.001 |
| PB Thickness vs Overlap | 0.22 | 0.10 | 0.34 | < 0.001 |

PB – peroneus brevis, PL – peroneus longus.

#### 2.8. Presence of low-lying peroneus brevis muscle belly

Low-lying muscle belly (LLMB) was most frequently observed in tendons overlapping the peroneus longus and protruding medially, with 60 of 161 cases (37%) demonstrating a low-lying muscle belly in this configuration. The presence of LLMB differed across peroneus brevis tendon positions, but this variation did not reach statistical significance (χ²(3) = 6.17, p = 0.105; Table 5).

**Table 5.** The presence of the low-lying peroneus brevis muscle belly.

| Relative position | No | Yes | Total |
| --- | --- | --- | --- |
| PB medial of PL | 26 | 7 | 33 |
| PB overlaps PL and protrudes laterally | 18 | 4 | 22 |
| PB overlaps PL and protrudes laterally and medially | 8 | 6 | 14 |
| PB overlaps PL and protrudes medially | 101 | 60 | 161 |
| Total | 153 | 77 | 230 |

PB – peroneus brevis, PL – peroneus longus.

#### 2.9. The presence of peroneus quartus

The peroneus quartus was observed in 18.3% of cases (Table 6). Fisher’s exact test revealed a significant association between the presence of the peroneus quartus and the relative position of the peroneus brevis tendon (p = 0.0124).

**Table 6.** Relative position of peroneus brevis and longus in relation to peroneus quartus.

| Relative position of the peroneal tendons | Absence (n) | Absence (%) | Presence (n) | Presence (%) |
| --- | --- | --- | --- | --- |
| PB medial of PL | 22 | 9.6% | 11 | 4.8% |
| PB overlaps PL and protrudes laterally | 16 | 7.0% | 6 | 2.6% |
| PB overlaps PL and protrudes laterally and medially | 10 | 4.3% | 4 | 1.7% |
| PB overlaps PL and protrudes medially | 140 | 60.9% | 21 | 9.1% |
| Total | 188 | 81.7% | 42 | 18.3% |

PB – peroneus brevis, PL – peroneus longus.

The highest percentages of the presence of the peroneus quartus were in the general flat and flattened convex laterally tendons (Table 7). Both the Chi-square test (X² = 6.00, p = 0.1114) and Fisher’s Exact Test (p = 0.1051) suggest that there is no significant association between the shape of the peroneus brevis and the presence of the peroneus quartus.

**Table 7.** The shape of the peroneus brevis on the cross-sectional area vs the presence of the peroneus quartus.

| Shape of Peroneus Brevis | Absence (n) | Absence (%) | Presence (n) | Presence (%) |
| --- | --- | --- | --- | --- |
| Flattened convex laterally | 17 | 7.4% | 8 | 3.5% |
| Flattened convex medially | 71 | 30.9% | 10 | 4.3% |
| General flat | 68 | 29.6% | 14 | 6.1% |
| Oval | 32 | 13.9% | 10 | 4.3% |
| Total | 188 | 81.7% | 42 | 18.3% |

#### 2.10. Modelling of tendon position

A multiple linear regression model was used to assess the association between variables and the peroneal tendons overlap in mm. Seventeen influential observations were excluded based on Cook’s distance (threshold: 4/n). The final model included four predictors: the presence of the peroneus quartus muscle, the presence of a low-lying muscle belly, the cross-sectional area of the peroneus longus, and the width of the peroneus brevis (Table 8, Figure 14). VIF values for all predictors ranged from 1.03 to 3.69, indicating no concerning multicollinearity.

**Figure 13.**
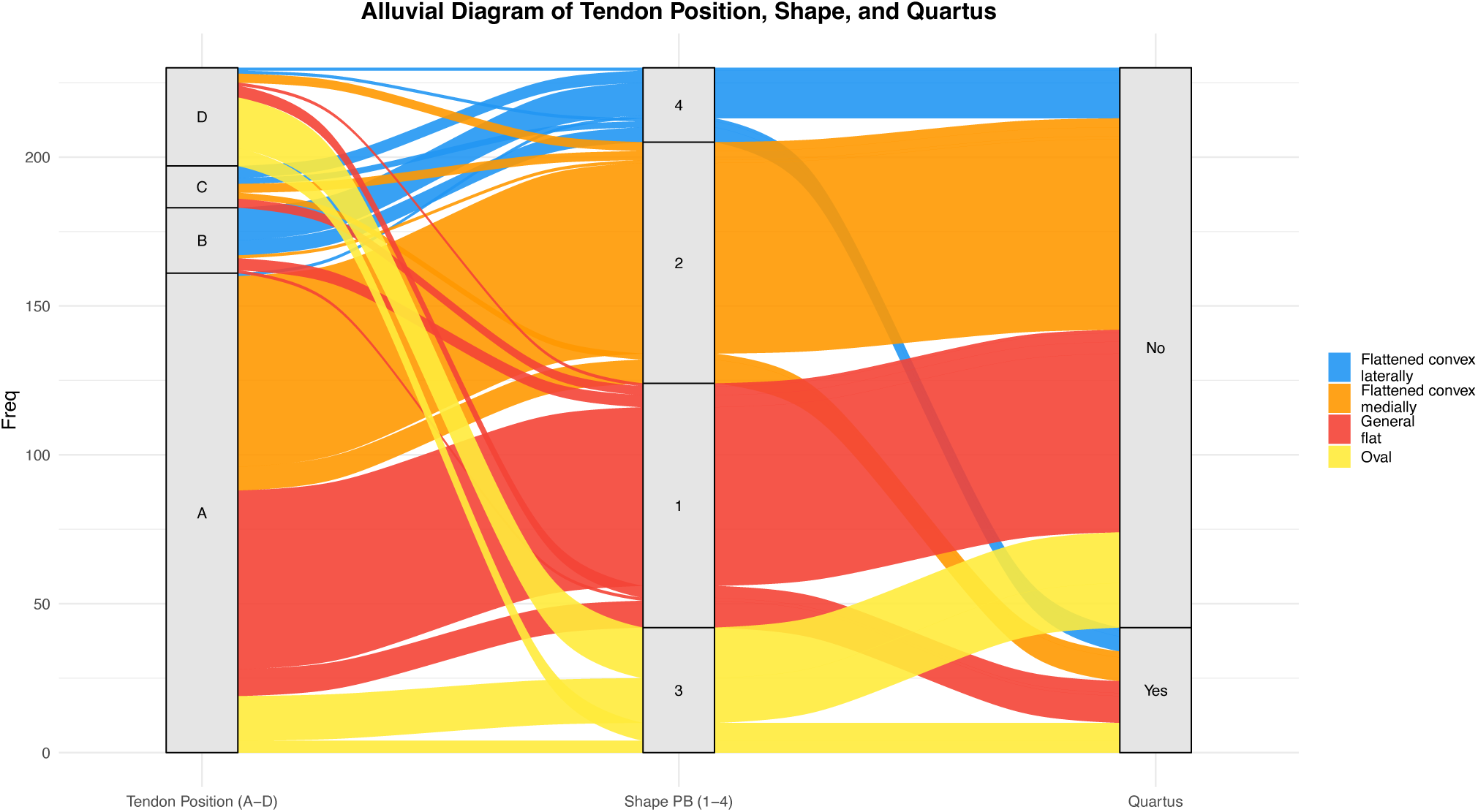
The alluvial diagram shows the relationship between the peroneus brevis (PB) position, the shape of the tendons on the transverse cross-section, and the presence of the peroneus quartus. Peroneus brevis positions are labelled A to D: A – PB overlaps the peroneus longus (PL) and protrudes medially, B – PB protrudes laterally, C – PB protrudes both laterally and medially, and D – PB is medial to PL. The PB shape is classified into four groups: 1– general flat, 2 – flattened convex medially, 3 – oval, and 4 – flattened convex laterally. Presence and absence of the peroneus quartus are marked as yes or no. The width of the links in the diagram indicates the distribution of tendons in each category.

**Figure 14.**
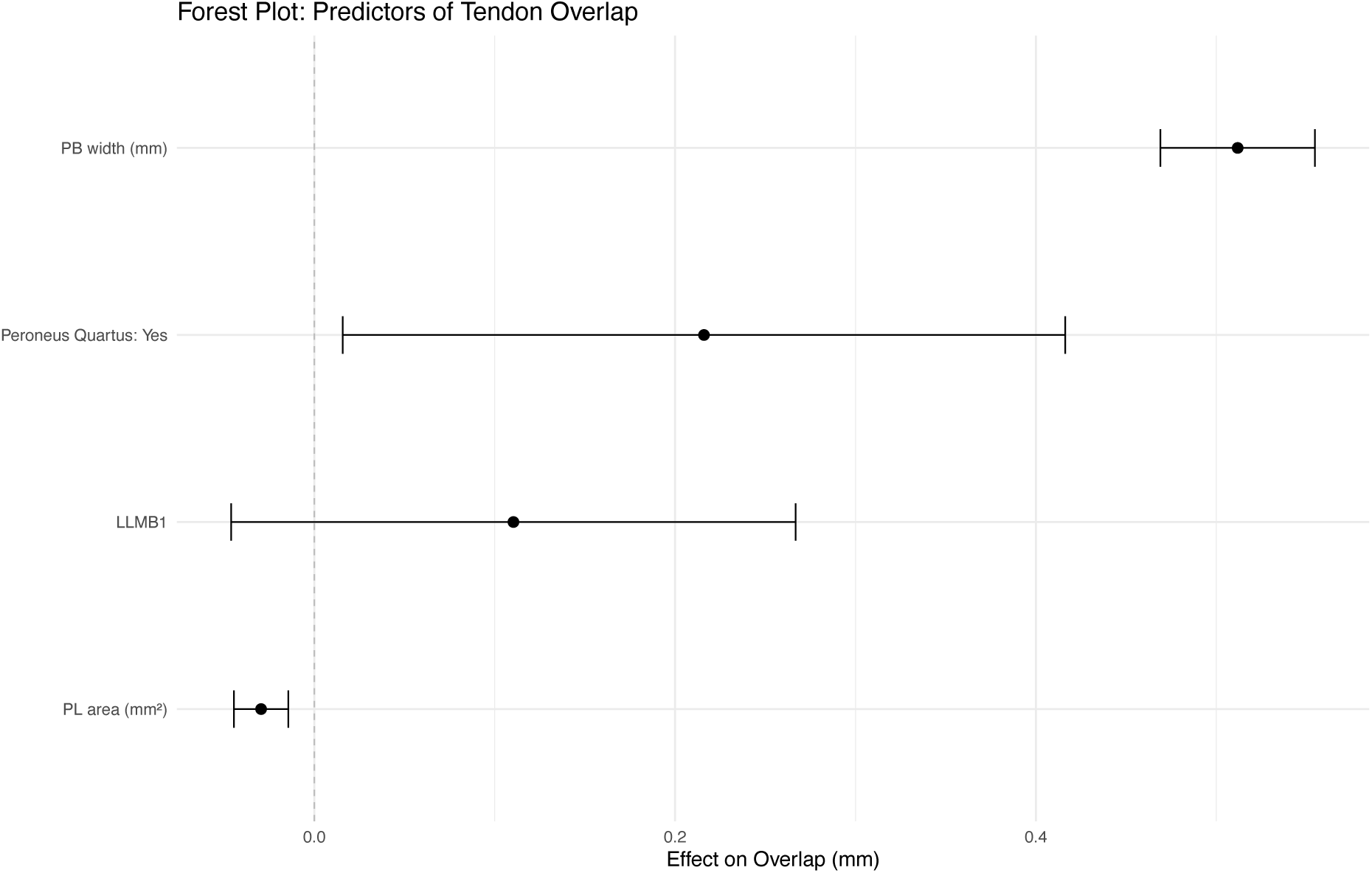
Forest plot showing effect sizes and 95% confidence intervals for predictors of tendon overlap. Peroneus brevis width had the strongest association, followed by peroneus longus area and presence of a peroneus quartus. LLMB1-presence of low-lying muscle belly of peroneus brevis. PB-peroneus brevis, PL-peroneus longus.

**Table 8.** Linear regression model evaluating predictors of peroneal tendon overlap (in millimetres) after exclusion of influential observations. Estimates represent unstandardised regression coefficients.

| Term | Estimate | Std error | Statistic | p value | conf low | conf high |
| --- | --- | --- | --- | --- | --- | --- |
| (Intercept) | -1.15 | 0.18 | -6.46 | 0.00 | -1.51 | -0.80 |
| Peroneus quartus | 0.22 | 0.10 | 2.13 | 0.03 | 0.02 | 0.42 |
| Low-lying muscle belly of peroneus brevis | 0.11 | 0.08 | 1.39 | 0.17 | -0.05 | 0.27 |
| Peroneus longus cross section area in mm<sup>2</sup> | -0.03 | 0.01 | -3.86 | 0.00 | -0.04 | -0.01 |
| Width of the peroneus brevis tendon in mm | 0.51 | 0.02 | 23.54 | 0.00 | 0.47 | 0.55 |

Moderate correlations were observed between peroneus brevis width and area (r = 0.67), and between brevis and longus area (r = 0.70), while all other correlations were weak (Supplementary Table S4). The presence of the peroneus quartus was significantly associated with increased tendon overlap (β = 0.22 mm, 95% CI: 0.02–0.42, *p* = 0.03). Although predicted values suggested a steeper increase in overlap among individuals with both a peroneus quartus and a low-lying muscle belly (Figure 15), the latter did not reach statistical significance in the multivariable model (*p* = 0.17). The overlap increased substantially with peroneus brevis width (β = 0.51 mm per mm, 95% CI: 0.47–0.55, *p* < 0.001) and decreased slightly with greater peroneus longus area (β = –0.03 mm per mm², 95% CI: –0.04 to –0.01, *p* < 0.001).

**Figure 15.**
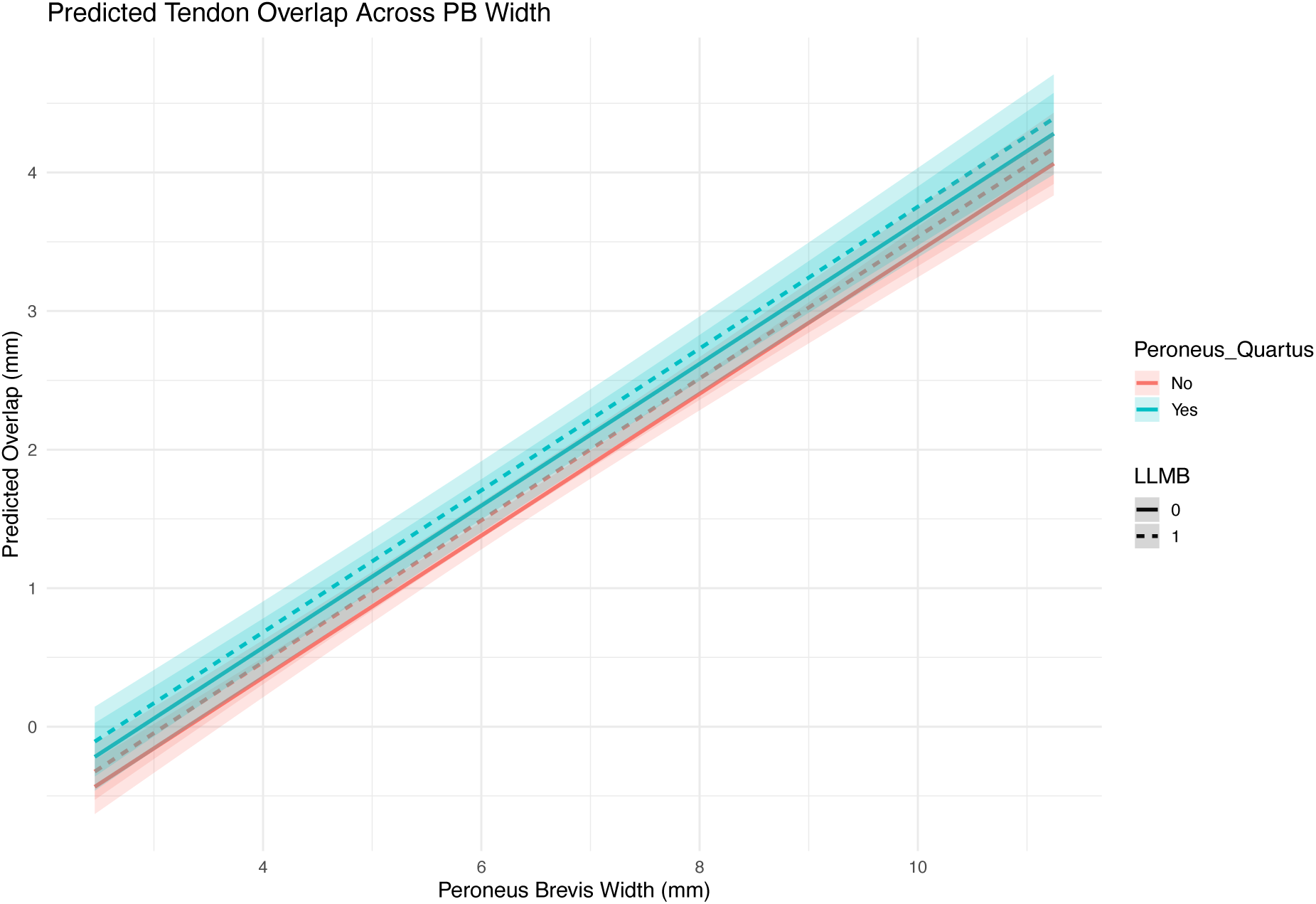
Predicted tendon overlap (mm) across the range of peroneus brevis widths, stratified by presence of a peroneus quartus and a low-lying muscle belly. Overlap increased consistently with width, with the steepest slope observed in cases with both anatomical variants. LLMB1-presence of low-lying muscle belly of peroneus brevis. PB-peroneus brevis.

Details on multicollinearity, influential observations, and robustness analyses (including standardised coefficients and bootstrapped confidence intervals) are available in the appendix (Tables S1–S5).

## Discussion

To the best of our knowledge, this is the first study to systematically assess anatomical variation in the position of the peroneus brevis tendon and factors that impact it in a large in vivo population with MRI. Our study demonstrates that the position and morphology of the peroneus brevis tendon exhibit substantial anatomical variation. In the most common configuration, the peroneus brevis overlaps the peroneus longus and protrudes medially, yet other variants were identified. Tendon shape was closely related to its position: medially positioned tendons were typically oval-shaped, while overlapping tendons, particularly those protruding laterally, were more often flattened and wider. Flattened tendons also had a larger cross-sectional area, indicating that tendon shape and size are strongly interconnected.

Furthermore, the presence of the peroneus quartus muscle was most often seen in tendons with lateral or combined protrusions, especially those with a laterally convex shape. These interrelated anatomical features point to a structured, rather than random, variation in tendon morphology. We noticed that tendons that protrude laterally in asymptomatic individuals may, in our opinion, be misinterpreted as instability since the peroneus brevis tend to be more lateral in relation to the peroneus longus. Interpreting this variant as pathology may lead to unnecessary surgery. To better understand relationships between position, we performed multivariate regression analysis to quantify the impact of specific anatomical features on tendon positioning.

The position between the peroneal tendons is particularly relevant in the context of tendon instability assessment and the possible risk of the split tear. Moreover, the cross-sectional area of the tendon was related to its shape, with flattened convex medially having the largest area. Flattened tendons, particularly those with lateral convexity, exhibited the largest cross-sectional areas, while oval tendons had the smallest. During dorsiflexion, the peroneus brevis tendon becomes more flattened and compressed against the fibular groove by the overlying peroneus longus tendon. This observation from the previous studies is true if the tendons are related in the most common variant. But if the peroneus brevis is located medially, this theory is not valid. However, there are no studies regarding which anatomical shape is more related to the split. Pathomechanics for longitudinal splitting is not known; some studies suggest tendon shape as a potentially important factor in the pathogenesis of split tears (Rademaker et al., 2000). Tendons positioned in overlapping configurations had significantly greater width and thickness, indicating that bulkier tendons are more likely to deviate from a colinear arrangement with the peroneus longus. It can be hypothesized that increased tendon cross-sectional area may mechanically drive or accommodate changes in position.

We observed that older individuals had larger peroneus brevis cross-sectional areas than younger patients, without associated signal abnormalities or changes in tendon morphology. This likely reflects physiological age-related changes rather than pathology, consistent with previous observations of tendon thickening in older populations (Bowden et al., 2018; Hodgson et al., 2012). Recognizing such anatomical variation is particularly important in ageing patients, where normal findings may be misinterpreted as tendinosis or split tear.

A multiple linear regression model confirmed that the presence of a peroneus quartus muscle and the width of the peroneus brevis tendon were independently associated with increased tendon overlap. Overlap also increased with a smaller peroneus longus area, while the low-lying muscle belly showed no independent effect. Notably, the predicted values suggested steeper increases in overlap among individuals with both anatomical variants, but only the peroneus quartus reached statistical significance. The dynamic balance between the components of the superior peroneal tunnel may be an important factor influencing the position of the peroneus brevis tendon. Findings from our study indicate that the peroneus brevis tendon position is variable and related to anatomical factors, such as its own morphology, peroneus longus, the presence of peroneus quartus and low-lying muscle belly. These variations should be recognized in clinical practice, especially in the evaluation of peroneal instability, pre-surgical planning, and in distinguishing anatomical variants from pathology. It is particularly important for radiologists reporting ankle MRI not to take the normal finding as pathology.

Presence of the peroneus quartus muscle may modify the relationship between structures. The peroneus quartus was most frequently observed in cases where the peroneus brevis tendon protruded laterally or medially, particularly when the tendon displayed lateral convexity. This suggests that accessory musculature may contribute to changes in tendon trajectory during flexion, possibly through altered biomechanics or competition for space in the superior peroneal tunnel. However, due to the limited number of quartus-positive cases, these associations should be interpreted cautiously, and further studies are needed.

Our study has several limitations, including its retrospective design, lack of histological correlation, and single-center setting. Future studies incorporating multicenter data and histological validation could provide further insights into the clinical significance of these anatomical variations.

## Conclusion

We identified four anatomical variants in the position of the peroneus brevis tendon relative to the peroneus longus on MRI: medial, and overlapping with medial, lateral, or combined protrusion. The most common was overlap with medial protrusion. Tendon position was significantly associated with shape, cross-sectional area, and the presence of a peroneus quartus muscle. In contrast, the presence of a low-lying muscle belly showed no independent association. These configurations represent normal anatomical variation. Recognizing them in clinical practice may help avoid misinterpreting normal findings as pathology. Future studies should assess whether localizations variants predispose to instability or tendon split tear.

## Declarations

### Ethics approval and consent to participate

All procedures performed in studies involving human participants were in accordance with the ethical standards of the institutional and/or national research committee and with the 1964 Helsinki Declaration and its later amendments or comparable ethical standards. The Swedish Ethical Authority approved this study and waived informed consent (number 2024-07283-02).

### Consent for publication

Not applicable. The manuscript does not contain the data of individuals in any form.

### Availability of data and material

Yes

### Competing interests

The authors declare that they have no competing interests.

### Funding

This work was supported by Stiftelsen Tornspiran (934: 2023-12-01) and by the Swedish state under the agreement between the Swedish Government and the county councils, the ALF-agreement (ALFGBG-1006741).

### Authors’ contributions

Conceptualization: PS; Methodology: PS, KBD, RZ; Software: PS, RZ; Data Collection: All authors; Data Curation: RZ, PS; Writing – Original Draft: RZ, PS; Writing – Review and Editing: All authors. All authors contributed to previous manuscript versions, and all have read and approved the final manuscript.

## Data Availability

All data produced in the present study are available upon reasonable request to the authors

## Acknowledgements

None

## Appendix

## Notes

### Competing Interest Statement

The authors have declared no competing interest.

### Funding Statement

This study was funded by Stiftelsen Tornspiran (934: 2023-12-01) and by the Swedish state under the agreement between the Swedish Government and the county councils, the ALF-agreement (ALFGBG-1006741).

### Author Declarations

The Swedish Ethical Authority approved gave ethical approval for this work (number 2024-07283-02).

### Summary of Updates

We updated sections methods, results, discussion – reformulated and shortened some sections.

